# Trust in Health Information Sources Among Patients with Systemic Lupus Erythematosus in the Social Networking Era: The TRUMP^2^-SLE Multicentre Study

**DOI:** 10.1101/2024.08.17.24312148

**Authors:** Takanori Ichikawa, Dai Kishida, Yasuhiro Shimojima, Nobuyuki Yajima, Nao Oguro, Ryusuke Yoshimi, Natsuki Sakurai, Chiharu Hidekawa, Ken-ei Sada, Yoshia Miyawaki, Keigo Hayashi, Kenta Shidahara, Yuichi Ishikawa, Yoshiki Sekijima, Noriaki Kurita

## Abstract

**Objectives:** The rise of social networking services (SNS) has significantly impacted how patients with systemic lupus erythematosus (SLE) acquire health information, potentially influencing their discussions with healthcare providers. This study aimed to identify the preferences, actual access, and trust levels in various health information sources among SLE patients, along with associated factors.

**Methods:** A cross-sectional study was conducted from June 2020 to August 2021 across five university medical centres in Japan, involving 510 SLE patients aged 20 years and older. The study measured access to and preferences for health information sources, including SNS, and assessed trust in these sources. Factors influencing, such as internet usage and health literacy (HL) (functional, communicative, and critical), were analyzed using Poisson regression with robust error variance.

**Results:** Among the respondents, 98.2% expressed trust in doctors, whereas lower trust was observed in websites/blogs (52.0%) and SNS (26.9%). Despite this, the internet was the most frequent initial source of health information (45.3%), encompassing medical institutions’ homepages, patient blogs, Twitter, and Instagram. Longer internet use was linked to increased trust in homepages/blogs and SNS. Higher functional HL correlated with greater trust in doctors and lower trust in websites/blogs and SNS, while higher communicative HL was associated with increased trust in doctors, homepages, and blogs.

**Conclusion:** Many SLE patients seek online health information, including SNS, before consulting rheumatologists. Internet usage duration and multidimensional HL influence trust in online health information sources. Rheumatologists and healthcare providers should account for these factors when disseminating health information and engaging with patients.

**Key messages:** What is already known on this topic

- Although online health information seeking prior to patient visits is common among patients with systemic lupus erythematosus, the actual information sources remain unclear in the social networking era.

What this study adds

- The most common information sources to be accessed were the Internet, with the most frequent being medical institutions’ websites, patients’ homepages or blogs, and a small but not negligible proportion of social networking services (SNS).
- While trust in doctors and other healthcare professionals was high, trust in homepages and SNS was relatively low.
- Longer Internet usage was associated with greater trust in homepages, blogs, and SNS, whereas higher functional health literacy was associated with lower trust in these sources.

How this study might affect research, practice or policy

- These findings highlight that varying levels of trust in these sources and are influenced by factors such as Internet duration and healthy literacy dimensions.
- These findings underscore the need for rheumatologists and healthcare workers to consider patients’ preferences and trust levels when providing health information and engaging in patient interactions.

## Introduction

The widespread use of the Internet (IT) and social networking services (SNS) [1,2] has drastically changed how patients with systemic lupus erythematosus (SLE) access medical information [3]. As a result, rheumatologists are increasingly encountering patients who prepare for their visits using online sources [3,4]. Studies show that 80% of SLE patients seek disease-related information online, [5] and over 70% of those with rheumatic diseases use SNS [6]. Despite this, the specific online sources that SLE patients rely on and the factors that influence their trust in these sources remain largely unknown.

Understanding the varying levels of trust in SLE patients place in different health information sources, along with the characteristics that influence this trust, is critical for effective patient communication [3,7]. While many SLE patients report trusting online information [5], including SNS [2], this trust may be misplaced due to the potential spread of misinformation and misunderstandings in the absence of professional guidance [5,7]. Previous research has identified factors such as age, [1,8] sex, [1,8] education, [8] and functional health literacy (HL) [9] that influence trust in the general population. However, it is unclear whether these predictors apply similarly to SLE patients, who may rely heavily on online information due to the chronic nature of their condition and the challenges of managing it.

Therefore, this study aims to investigate the actual use of health information sources, the degree of trust in these sources–particularly online media–and the patient characteristics that influence this trust, using data from the Trust Measurement for Physicians and Patients with SLE (TRUMP^2^-SLE), a multicentre study involving Japanese adult SLE patients.

## Methods

### Study design and participants

This cross-sectional study utilized baseline data from the TRUMP^2^-SLE study, a multicentre cohort study conducted at five academic medical centres: Showa University Hospital, Okayama University Hospital, Shinshu University Hospital, Yokohama City University Hospital, and Yokohama City University Medical Centre. The study adhered to the Declaration of Helsinki and Good Clinical Practice guidelines and was approved by the Ethics Review Board of Shinshu University (approval number 5433). Participants included SLE patients aged 20 years or older, diagnosed according to the revised 1997 American College of Rheumatology classification criteria, who were receiving rheumatology care at the participating centres. Patients had to be able to complete the questionnaire survey independently. Informed consent was obtained from all participants prior to their enrolment in the study. Exclusion criteria included dementia or total blindness.

### Data collection

The questionnaire was administered at each facility between June 2020 and August 2021. Participants were asked to complete the survey either in the waiting room or at home. The questionnaire assured participants that their responses would remain confidential and would not be accessible to their attending physicians, with data being used exclusively for aggregation at the central facility.

### Health information sources: Preferred vs. actual access

To assess patients’ preferred and actual sources of health information, we adapted items from the Health Information National Trends Survey (HINTS), a national survey of the U.S. public’s use of and attitudes toward health-related information. [1,10] Patients were asked to identify their preferred source of health information by responding to the prompt: “Imagine you have a strong need to get information about your disease or treatment”. They could select one option from the following: books; pamphlets; doctor; healthcare provider other than a doctor, family/friend, television/radio, doctor’s lecture, newspapers/magazines; official patient groups (e.g., National Friendship Association for Collagen Disease), internet (homepage, blog), YouTube, Twitter, Facebook, Instagram, e-mail lists, and messaging applications (LINE, Facebook Messenger, etc.).

To determine the actual source of health information accessed, participants were then asked: “The most recent time you looked for information about your disease or treatment, where did you actually go first?" If an online source was chosen, participants were asked to specify the type of online information source (multiple selections allowed; Supplementary Items).

### Trust in Health Information Sources

Patient trust in health information sources was assessed by modifying the aforementioned survey items. Participants responded to the question: “How much do you generally trust the information about your disease and treatment obtained from each of the following sources?” [1,10] with four possible answers: “a lot of trust”, “some trust”, “neither trust nor distrust”, and “distrust”. The sources evaluated included books; pamphlets; doctor; healthcare provider other than a doctor, family/friend, television/radio, doctor’s lecture, newspapers or magazines, official patient groups (such as National Friendship Association for Collagen Disease); IT (homepage, blog); SNS (Twitter, Facebook, etc.); and patient group messaging applications (LINE, Facebook messenger, etc.)

### Measurement of covariates

Covariates included variables that were potential confounders influencing both health information sources and trust in those sources, as identified in existing literature. These variables included age, [1] sex, [1] education, [1] household income, [1] marital status, health literacy (HL), [11] IT usage time per day, [11,12] disease duration, [13] disease activity, organ damage, history of disease flare-ups, experience of adverse drug events, and emotional health.

Multidimensional HL was measured using the validated 14-item Functional Communicative Critical Health Literacy Scale (FCCHL) [14], which assesses three domains: functional HL (ability to read and understand healthcare instructions), communicative HL (ability to extract and communicate health information), and critical HL (ability to critically analyse and use health information). Each item was scored on a 4-point scale, with domain scores ranging from 1 (low HL) to 4 (high HL).

IT usage time per day was assessed by asking participants: "How much time do you spend using the Internet and social networking services daily, not including work time?" [12] with respondents categorized into four levels: "none”, "less than 1 hr”, "1 to 2 hrs”, and "more than 2 hrs".

Disease activity was evaluated by the attending physician using the Systemic Lupus Erythematosus Disease Activity Index 2000 (SLEDAI-2K). Organ damage was assessed using the Systemic Lupus International Collaborating Clinic Damage Index (SDI). A history of disease flare-ups was defined as any clinically evident worsening of symptoms in one or more organs.

Adverse effects from lupus medications were measured with the single item: “Lupus medication(s) bothersome side effects”, based on the Japanese version of Lupus PRO, with participants responding according to their experiences over the past four weeks [15,16].

Emotional health was measured using a single domain of the Lupus-PRO, consisting of six items scored on a 4-point scale, [15] with the total score converted to a scale ranging from 0 to 100.

### Statistical analysis

All statistical analyses were conducted using Stata/SE, version 16.1 (StataCorp, College Station, TX, USA). Categorical variables were described as frequencies and proportions, while continuous variables were summarized as medians and interquartile ranges (IQR). Bar graphs illustrated the proportions of patients’ preferred and actual health information sources. IT access was defined as first access to any of the following sources: homepage/blog, YouTube, Twitter, Facebook, Instagram, e-mail lists, and messaging applications (LINE, Facebook messenger, etc.). For respondents who chose an online source, specific online sources selected were shown in bar graphs. Trust levels in health-information sources were depicted in stacked bar graphs. To analyse factors associated with trust in health information sources, we focused on three sources: doctors, IT (homepage, blog), and SNS (Twitter, Facebook, etc.) A modified Poisson regression model, adjusted for age, sex, education, household income, marital status, health literacy (HL), experience of adverse drug events, IT usage time, emotional health, disease activity, organ damage, disease flare-ups, and illness duration., was used to estimate the relative risk (prevalence ratio [PR] of factors influencing trust. The modified Poisson regression was chosen to address the overestimation of odds ratios when trust levels are not rare. [17] Missing covariates were handled using multiple imputations with chained equations, under the assumption that the data were missing at random. [18] Statistical significance was set at P < 0.05.

## Results

### Patient characteristics

Among the 516 SLE patients enrolled in the TRUMP^2^-SLE study, six were excluded due to incomplete or inconsistent responses to key questions about their health information sources and trust levels. Therefore, 510 patients were included in the final analysis. The participant characteristics are shown in Table 1.

**Table 1.** Patient characteristics (N= 510)

|  | **Total**<br>N = 510 |
| --- | --- |
| **<u>Demographics</u>** |  |
| Age, yr | 45.4 [36—55.6] |
| Women, n (%) | 450 (88.2 %) |
| missing, n | 0 |
| Education, n (%) |  |
| Junior High school or lower | 22 (4.6 %) |
| High school/College | 325 (68.3 %) |
| University/Graduate school | 129 (27.1 %) |
| missing, n | 34 |
| Household income, n (%) |  |
| <1,000,000 yen | 36 (8.5 %) |
| 1,000,000 to <5,000,000 yen | 185 (43.4 %) |
| 5,000,000 to <10,000,000 yen | 162 (38 %) |
| >10,000,000 yen | 43 (10.1 %) |
| *missing, n* | 84 |
| Marital Status, n (%) |  |
| Married | 274 (56.4 %) |
| Divorced/Widowed | 44 (9.1 %) |
| Unmarried | 168 (34.6 %) |
| *missing, n* | 24 |
| Disease duration, yr | 12.4 [6.5 – 19.7] |
| *missing, n* | 9 |
| SLEDAI-2K, pts | 4 [2 – 6] |
| SDI, pts | 0 [0 – 1] |
| History of disease flare-up, n (%) | 69 (14.6 %) |
| *missing, n* | 38 |
| Experience of adverse drug event within the past month, n (%) | 150 (34.4 %) |
| *missing, n* | 74 |
| Multidimensional HL |  |
| Functional HL, pts | 3.6 [3.1 – 4] |
| *missing, n* | 10 |
| Communicative HL, pts | 3 [2.6 – 3.4] |
| *missing, n* | 10 |
| Critical HL, pts | 2.8 [2.3 – 3] |
| *missing, n* | 9 |
| Time used for IT per day, n (%) |  |
| None | 63 (12.4 %) |
| <1hr | 182 (35.9 %) |
| 1-<2hr | 127 (25.1 %) |
| 2hr< | 135 (26.6 %) |
| *missing, n* | 3 |
| Emotional health, pts | 75 [50 – 87.5] |
| *missing, n* | 71 |

<sup>a</sup>Median (interquartile range) are presented for continuous data
Column percentages were presented for categorical data
SLEDAI-2K: Systemic Lupus Erythematosus Disease Activity Index 2000; SDI: Systemic lupus International Collaborative Clinics Damage Index; IT: Internet; HL: Health literacy.

The median age of the patients was 45.4 years (IQR 36.0-55.6), and the majority (88.2%) were female. The median duration of SLE was 12.4 years (IQR 6.5-19.7), with a median SLEDAI-2K score of 4.0 (IQR 2.0-6.0), indicating moderate disease activity, and a median SDI score of 0 (IQR 0.0-1.0), reflecting minimal organ damage. In terms of HL, the median functional HL was 3.6 (IQR 3.1-4.0), communicative HL was 3.0 (IQR 2.6-3.4), and critical HL was 2.8 (IQR 2.3-3.0).

### Health information sources

Figure 1 illustrates the preferred and actual first sources of health information accessed by patients. While 66.3% (95%CI 61.9–70.5%) of patients indicated a preference for accessing physicians first, IT sources were the most commonly accessed in practice, with 45.3% (95% CI 40.8–49.8%) of patients turning to IT first. This was followed closely by physicians, with 41.6% (95% CI 37.2–46.1%) of patients actually consulting them first. The most frequently accessed IT sources were medical institution websites (69.6%, 95%CI 63.2– 75.6%), followed by patient homepages/blogs (39.7%, 95%CI 33.3–46.5%) and pharmaceutical company websites (34.4%, 95%CI 28.2–41.0%). SNS such as Twitter and Instagram, primarily posted by other patients rather than healthcare professionals, were also accessed. Among patient-sourced media, the most accessed platforms were patient websites/blogs, patient group websites (17.0%, 95% CI 12.3–22.5%), Twitter (8.5%, 95% CI 5.2–12.9%), and Instagram (6.3%, 95% CI 3.5–10.3%).

**Figure 1.**
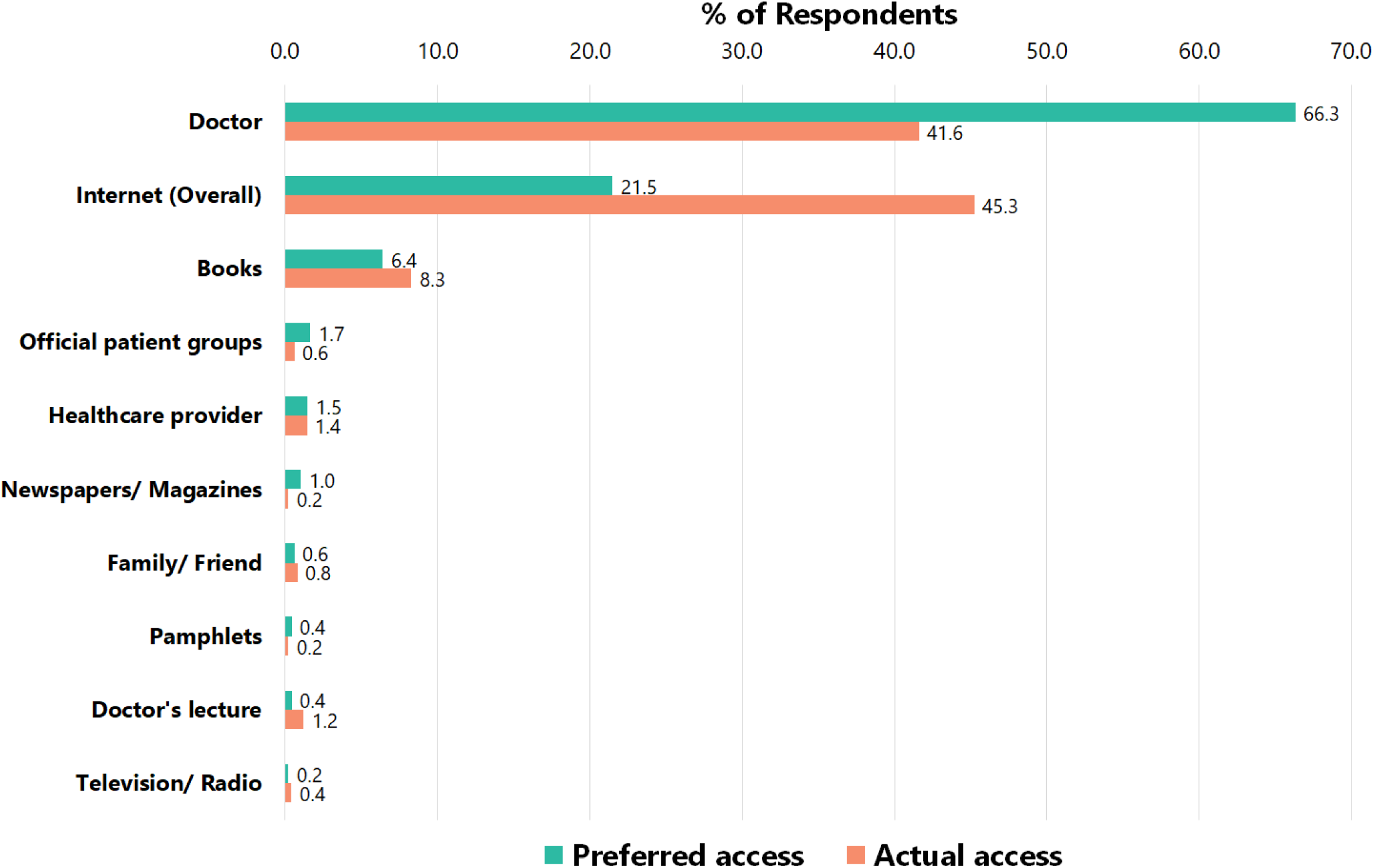
Healthcare information source patients would like to access first (n=484) and actually access first (n=495) IT was counted if one of the following was chosen: homepage/blog, YouTube, Twitter, Facebook, Instagram, e-mail list, or messaging applications (LINE, Facebook messenger, etc.). However, neither e-mail lists nor messaging applications have been used.

**Figure 2.**
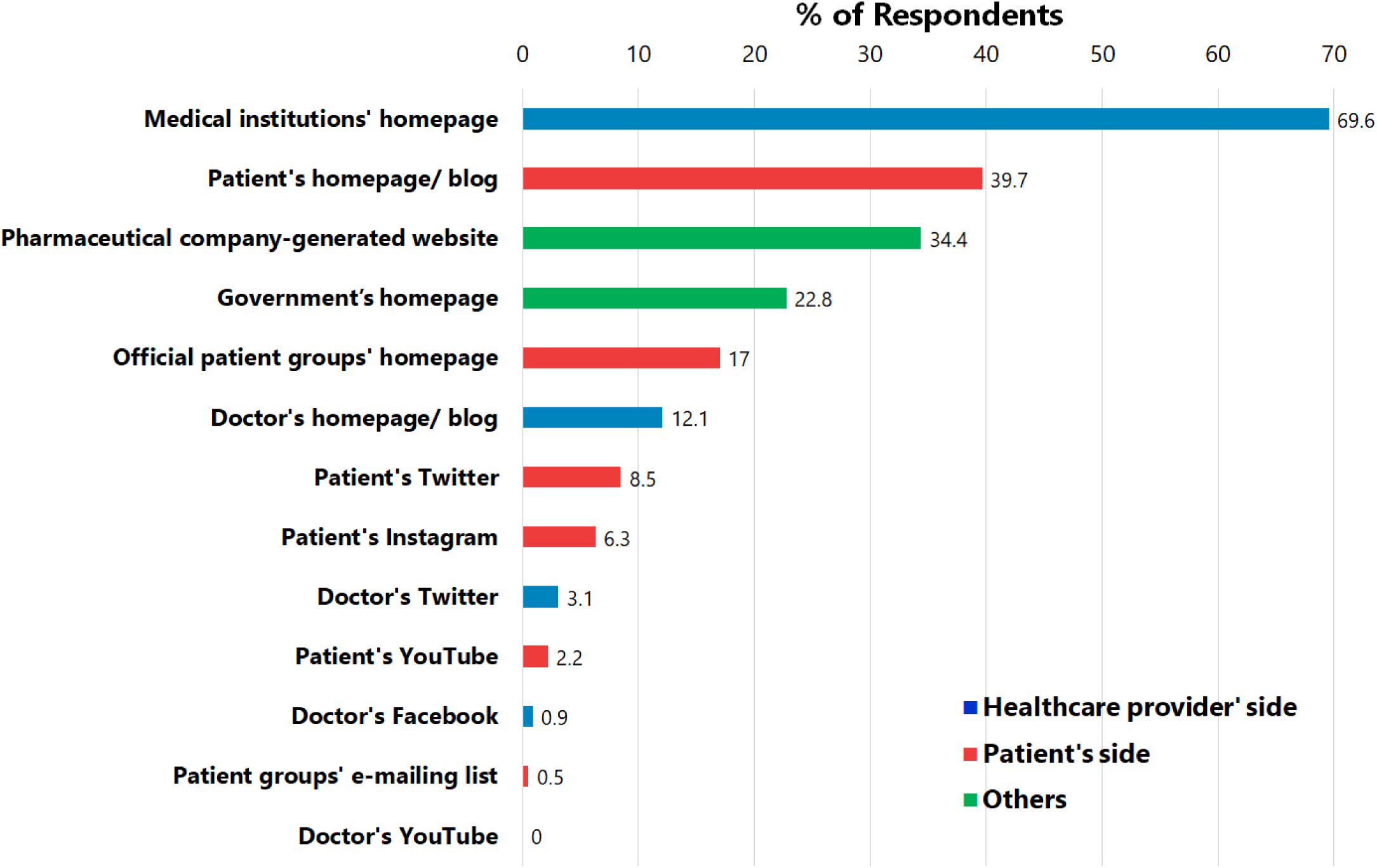
Internet health information sources actually accessed by patients (multiple choice, n = 224)

### Trust in health information sources

Figure 3 presents the levels of trust in various health information sources. Doctors were the most trusted source, with 98.2% (95% CI 96.6–99.2%) of patients expressing high trust, followed by other healthcare professionals (75.9%, 95% CI 71.8–79.6%) and doctor’s lecture (74.0%, 95% CI 69.9–77.9%). In contrast, trust in media sources was significantly lower, with television/radio trusted by 42.4 % (95% CI 38.0–46.8%) of patients. Trust in IT media was moderate, with websites/blogs trusted by 52.0% (95% CI 47.50–56.6%), but trust in patient group messaging applications (28.1%, 95% CI 24.1–32.3%) and SNS (26.9%, 95% CI 23.0–31.0%) was notably lower.

**Figure 3.**
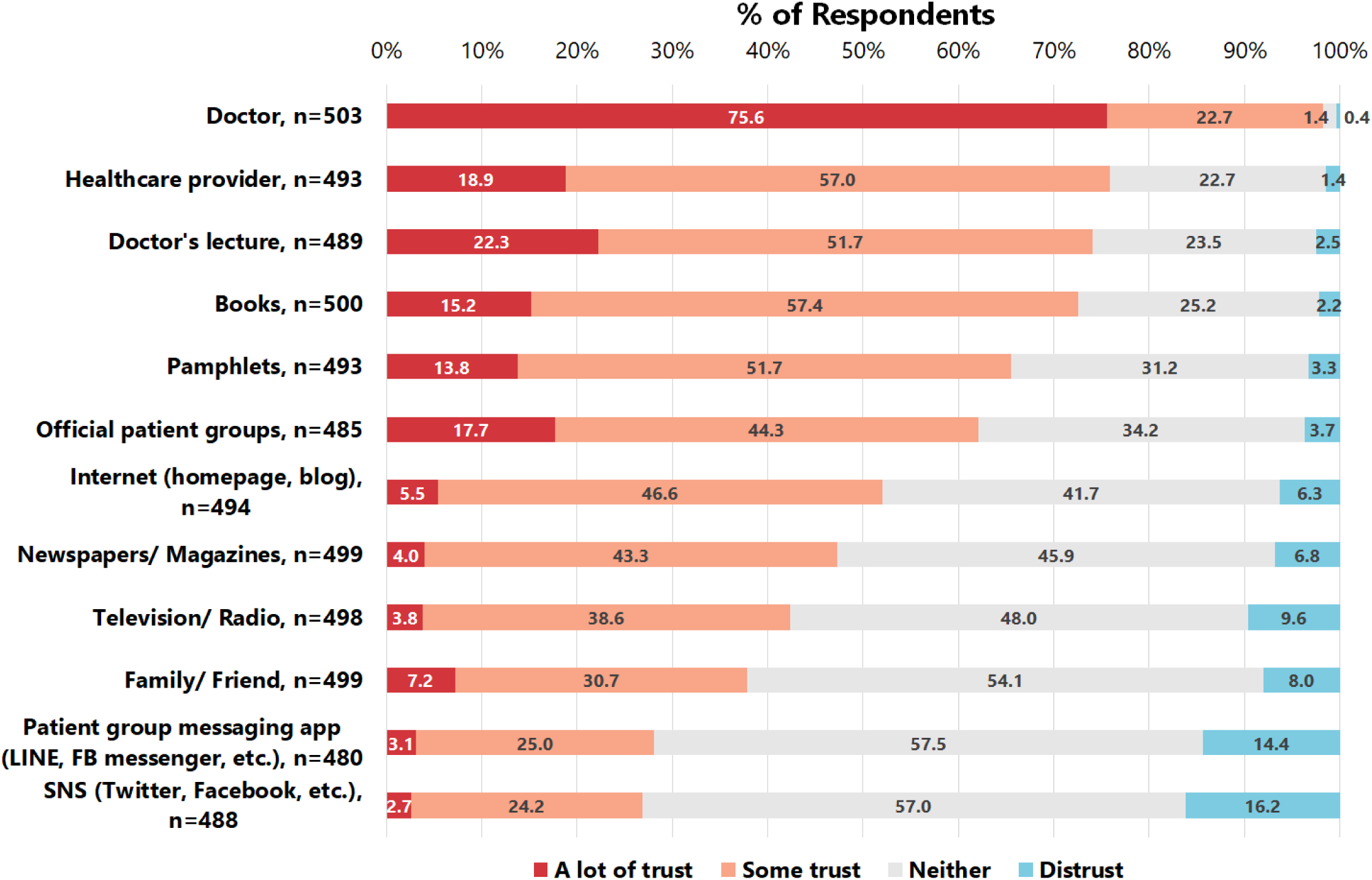
Level of trust in specific health information sources. n indicates number of respondents for each health information source. FB: Facebook

The analysis also revealed that longer IT usage was associated with higher trust in homepages/blogs and SNS (Table 2). Additionally, higher functional HL was associated with higher trust in physicians (adjusted PR [aPR] 1.14 [95% CI 1.02–1.27]) but lower trust in homepages/blogs and SNS (aPR 0.84 [95% CI 0.72–0.99] and 0.73 [95% CI 0.56–0.94], respectively). Higher communicative HL was associated with increased trust in both physicians (aPR 1.17 [95% CI 1.05 to 1.31] and homepages/blogs 1.32 [95% CI 1.09 to 1.61]).

**Table 2.** Patient characteristics associated with trust in respective health information sources.

|  | Doctor (n =503) | Internet (homepage/ blog) (n =494) | SNS (Twitter, Facebook, etc.) (n =488) |
| --- | --- | --- | --- |
|  | aPR, point estimate (95%CI) | aPR, point estimate (95%CI) | aPR, point estimate (95%CI) |
| Multidimensional health literacy |  |  |  |
| Functional HL, per 1-pt higher | **1.14 (1.02 to 1.27), p = 0.02** | **0.84 (0.72 to 0.99), p = 0.033** | **0.73 (0.56 to 0.94), p = 0.014** |
| Communicative HL, per 1-pt higher | **1.17 (1.05 to 1.31), p = 0.004** | **1.32 (1.09 to 1.61), p = 0.005** | 1.31 (0.93 to 1.83), p = 0.123 |
| Critical HL, per 1-pt higher | 0.9 (0.81 to 1), p = 0.061 | 0.89 (0.74 to 1.07), p = 0.213 | 0.84 (0.62 to 1.15), p = 0.286 |
| Duration of internet use |  |  |  |
| None | Reference | Reference | Reference |
| <1 h | 1.01 (0.85 to 1.2), p = 0.882 | **3.48 (1.82 to 6.64), p <0.001** | **3.7 (1.4 to 9.76), p = 0.008** |
| 1 h-<2 h | 0.94 (0.76 to 1.15), p = 0.534 | **4.26 (2.2 to 8.27), p <0.001** | **4.35 (1.62 to 11.69), p = 0.004** |
| >2 yr | 1 (0.8 to 1.24), p = 0.97 | **4.18 (2.11 to 8.27), p <0.001** | **4.28 (1.54 to 11.9), p = 0.005** |

The outcome measure was trust in each health information source. Trust was considered present if participants reported having “a lot” or “some” trust. An exception was made for trust in doctors, which was considered only if “high trust” was reported. This distinction was necessary because if both “a lot of trust” and “some trust” were counted as trust, most patients would be categorized as having trust, making the analysis less meaningful. To estimate the prevalence ratio for trust in relation to patient characteristics, a modified Poisson regression (Poisson regression with robust error variance) was used. The regression model included the following explanatory variables: age; sex; education; household income; marital status; disease duration; multidimensional health literacy; time spent on IT; SLEDAI; SDI; emotional health as measured by LupusPRO; disease flare-ups; and a history of adverse drug events in the past month.
aPR, adjusted prevalence ratio; CI, confidence interval; SNS, social networking service; HL, health literacy

## Discussion

### Preferred health information source and actual source patients accessed

Among adult patients with SLE, doctors were the most preferred health information source (66.3%), while IT was the most commonly accessed actual source (45.3%). Medical institution websites were chosen by approximately 70% of respondents, with patients’ homepages/blogs being the next most common source. A smaller but non-negligible proportion of patients (< 10%) utilized SNS.

Our findings contrast with previous reports. For instance, a small-scale study conducted during the acute phase of SLE [19] and another conducted before and after the COVID-19 pandemic [2] reported higher use of homepages (77%) [19] and SNS (approximately 30%), respectively. [2] These discrepancies likely stem from differences in survey timing and sampling methods. Previous studies recruited participants via online platforms, [19] e-mail, and SNS [2], resulting in a sample of regular IT users, potentially overestimating IT use in the general outpatient population. In contrast, our study, which involved a broader, more diverse patient population invited at their attending medical institutions, provides a more accurate reflection of actual health information-seeking behaviours, especially among patients in the maintenance phase with established treatment regimens. Importantly, 12.4% of our participants were infrequent IT users, underscoring the study’s broader applicability.

Our findings show that although doctors were preferred information sources, the Internet was more frequently used as the first information source by patients. This trend is consistent with the HINTS study conducted 20 years ago, which examined cancer-related information seeking. [1] Patients may use IT "to learn”, [7] gathering information in advance about managing their SLE [12] or understanding changes in symptoms or test results [5] during their outpatient visits to a rheumatologist every one to three months.

Our survey extended to the frequency of online information source access by senders. It highlighted that patients frequently accessed patient homepages/blogs (39.7%) and patient group websites (17.0%). Additionally, there was notable access to SNSs, including patients’ Twitter (8.5%), and Instagram (6.3%), as well as doctors’ Twitter. Patients’ access to health information sources, including homepages and SNS, may reflect their desire to learn from others with SLE, [5] connect with fellow patients, [5,7] share experiences, [7] and engage with rheumatologists [6]–potentially driven by feelings of loneliness. [5,20]

Based on these findings, rheumatologists and healthcare workers can take several actions. First, rheumatologists should inquire about the sources and senders of health information that patients discuss. [3] If a patient’s anxiety is heightened by misinformation from other patients’ SNS, [7] addressing the facts can help clarify the issue. [3,12,21] In addition, rheumatologists can guide patients toward high-quality, reliable sources, such as medical institutions, government homepages, or unbiased pharmaceutical company websites. Understanding patients’ connection-seeking behaviour on SNS can also allow rheumatologists to show empathy and help alleviate loneliness. Second, healthcare workers can strategically deliver high-quality, reliable information via SNS platforms such as Twitter, Instagram, and Facebook. [6] These channels are particularly important for young adult patients who prefer SNS as a source of accurate health information.

### Level of trust in specific health information sources and its correlates

High trust in doctors and other healthcare professionals was observed, while trust in homepages and SNS was relatively low. In addition, longer IT usage was associated with a greater likelihood of trusting homepages/blogs and SNS, whereas high functional HL was linked to lower trust in these sources.

Our study found relatively low trust rates compared to the very high trust in IT reported among SLE patients seeking health information (95%) [5] and the modest trust in SNSs (∼30%). [2] This difference may result from self-selection bias in previous studies [2,5]. The previous report of high trust in IT [5] supports our finding that longer IT use is linked to greater trust in IT. Similarly, our finding that longer IT use is linked to greater trust in SNS is supported by a previous study that reported increased SNS access was associated with trust in SNS. [2]

As expected, higher functional HL was linked to greater trust in doctors and lower trust in homepages/blogs and SNS. Our previous study also reported higher functional HL was associated with higher trust in their rheumatologists. [11] The lower trust in online information associated with higher functional HL may reflect patients’ experiences that such information is not always reliable [21] and that reading and wiring proficiency is needed to assess its accuracy. The greater trust in doctors and homepages/blogs linked to higher communicative HL may reflect SLE patients’ ability to extract and adapt information from these sources to their changing situations. [11]

Our findings offer several insights for rheumatologists and healthcare workers in their information provision practices. First, rheumatologists should communicate health information in plain language to build credibility, whether in person or via social media. [8,11] Second, when non-physicians provide information on homepages or SNS, prior supervision by a rheumatologist can help ensure accuracy, particularly for patients with low HL. [5] Third, using highly visible infographic formats can improve understanding and recall, especially for patients with low HL and limited IT experience. [22]

### Strengths and Limitations

This study has several strengths. First, the findings are generalizable to a broader population, as they were derived from multiple institutions in metropolitan and rural areas. Second, this study is the first to identify predictors of trust in specific health information sources, including SNS, while controlling various underlying characteristics such as age, disease activity, education level, and economic status.

However, there are some limitations to consider. First, the cross-sectional design of the study raises the possibility of reverse causation–for example, patients may use IT more extensively because they already trust the sources they access. Second, while the frequency of access by channel and sender was analysed, we could not assess the level of trust for each specific source. For instance, we could not differentiate the trust levels for SNS based on whether the sender was a patient or a rheumatologist. Finally, this study was conducted before the widespread adoption of large language models like ChatGPT in Japan; therefore, we could not assess how often patients with SLE use such language tools or how much they trust them. [12]

## Conclusion

In conclusion, this study elucidates the preferences of SLE patients regarding health information sources and their actual access patterns. It was observed that many patients turn to online resources, including patient homepages, SNSs, and healthcare provider websites, to gather information. Additionally, the study highlights the varying levels of trust in these sources, influenced by factors such as IT usage duration and health literacy dimensions. These findings underscore the need for rheumatologists and healthcare workers to consider patients’ preferences and trust levels when providing health information and engaging in patient interactions. Future research should explore how emerging technologies, such as large language models, may further shape these dynamics in the context of SLE.

## CONTRIBUTORSHIP STATEMENT

TI, DK, Y. Shimojima, NY, and NK designed the study. TI, DK, Y. Shimojima, NY, NO, RY, NS, CH, K. Sada, YM, KH and K. Shidahara were involved in patient recruitment and data collection. TI, DK, Y. Shimojima, NY, Y. Sekijima, and NK participated in the analysis. TI prepared the initial draft, and all authors were involved in reviewing the manuscript and providing critical comments.

## Data Availability

The datasets generated and/or analysed during the study are available from the corresponding author upon reasonable request.

## ACKNOWLEDGEMENTS

We thank Hiroko Nagasato, Kumi Sasaki, Yukari Hosaka (Showa University) and Miyuki Sato (Fukushima Medical University) for their assistance. The abbreviated name of the study, “TRUMP^2^-SLE (the Trust Measurement for Physicians and Patients with SLE)”, does not refer to a specific individual; instead, the name intends to suggest a trusted physician; an old meaning of the word “trump” is “a dependable and exemplary person”.

## Supplementary Item. Detailed question on actual accesses to online health information sources

Please answer the following question if you chose j (Internet [homepage/blog]) to p (messaging applications [such as LINE, Facebook messenger]) in the previous question. Which of the following sources did you use? Please **circle <u>all</u>** that apply.

a. Government’s homepage
b. Medical institutions’ homepage
c. Pharmaceutical company-generated website (e.g., SLE.jp, The LUPUS)
d. Official patient groups’ homepage (e.g., National Friendship Association for Collagen Disease)
e. Doctor’s homepage/blog
f. Doctor’s Twitter
g. Doctor’s Facebook
h. Doctor’s YouTube
i. Patient’s homepage/blog
j. Patient’s Twitter
k. Patient’s YouTube
l. Patient’s Instagram
m. Patient groups’ e-mailing list

